# Vasculopathy as a Mechanical Barrier to Cancer Spread: Clinical Evidence and a Rheology-Based Model in Lung Cancer

**DOI:** 10.64898/2026.01.12.26343968

**Authors:** Giulia M. Stella, Cristina Novy, Francesco Rocco Bertuccio, Ilaria Ferrarotti, Valentina Conio, Chandra Bortolotto, Tiziana Giorgiani, Lucrezia Pisanu, Ilaria Salzillo, Annalisa De Silvestri, Vittorio Arici, Alice Maccarini, Pietro Cerveri, Angelo G. Corsico, Antonio Bozzani

## Abstract

Metastatic dissemination in lung cancer (LC) and other solid tumors is influenced not only by tumor-intrinsic biology and immune–inflammatory responses, but also by the physical properties of the vascular system through which circulating tumor cells (CTCs) migrate. Peripheral arterial disease (PAD), particularly when manifesting as aneurysmal dilation, is frequent among long-term smokers and is associated with chronic vascular inflammation and altered hemodynamics. We hypothesized that PAD-related vascular remodeling and rheological alterations may influence tumor metastatic capacity. Through a retrospective analysis of 976 patients diagnosed with both cancer and arteriopathy between 2018 and 2024, a cohort of 120 individuals with concomitant aneurysmal and neoplastic disease was identified. Demographic, biochemical, and pathological variables were examined, and metastatic burden at diagnosis was compared with that of an unselected LC population from the same institution and with literature-reported data. We focused on non-small cell lung cancer (NSCLC) as a well-characterized biological model and developed a phenomenological biophysical framework linking inflammation-driven changes in blood viscosity to metastatic competence. A Monte Carlo simulation approach was used to estimate metastasis probability under control and PAD-like rheological conditions. Despite marked male predominance and high smoking exposure, the study cohort exhibited an unexpectedly low metastatic burden, with 13.3% of patients presenting metastatic disease at diagnosis and only 7.6% showing extrathoracic dissemination, compared with an expected rate of approximately 30%. Partition analysis identified arteriopathy as the strongest predictor associated with reduced metastatic dissemination. The rheological model indicated that once inflammation exceeds a critical threshold, increased blood viscosity and disturbed flow patterns may act as a mechanical filter impairing CTC extravasation. Monte Carlo simulations supported this threshold-dependent mechanism, showing an approximately 50% reduction in predicted metastatic rates in PAD-like conditions compared with controls. Collectively, these findings suggest that chronic PAD and aneurysmal vasculopathy may reshape the circulatory microenvironment, with NSCLC providing a mechanistically interpretable framework for a transition from a metastasis-permissive to a metastasis-restrictive rheological regime.

## 1. Introduction

Lung cancer (LC) frequently presents with distant metastases at diagnosis occurring in approximately 30% of cases [1, 2, 3]. Metastatic dissemination greatly affects both survival and quality of life. Indeed, resection of the primary tumor improves survival even in case of single metastases with preferential site of growth in the brain or bones rather than in the liver or lung itself [4, 5]. Moreover, the recent introduction of peri- and post-operative targeted therapies and immunotherapy has significantly improved outcomes LC patients, highlighting the crucial role of immune-inflammatory cascade in cancer progression [6, 7, 8]. It is, thus, mandatory to determine the biological features leading to LC early spreading. The latter is a highly complex and multi-step process encompassing the aberrant activation of multiple genetic and biologic processes, including epithelial-to-mesenchymal transition (EMT), angiogenesis and lymph angiogenesis. These events involve a dynamic crosstalk between the primary tumor mass, its stem cell compartment, and the surrounding microenvironment [9, 10, 11]. Once activated, cancer cell invasiveness relies on the induction of blood and lymphatic vessels, and on the cell ability to extravasate and reach distant organs [12]. Although extensive literature describes the molecular and genetic drivers of distant spreading, far fewer data are available about the complex interplay between invasive clones and the microenvironment through which they migrate, and on how this context ultimately shapes their fate. Studying cancer by focusing on blood viscosity and cell-flow interactions represent an innovative and realistic approach that may overcome some of the challenges the scientific community faces with traditional diagnostic methods such as imaging or sequencing. From this perspective, we reasoned that an altered vascular context could play a role in modulating cancer invasive properties. We performed first a retrospective clinical analysis to evaluate how changes in blood flow, in which metastatic cells circulate, and how altered biomechanics and hemodynamics resulting from arteriopathy, particularly aneurysmal dilation, a condition frequently found in smoker patients with lung cancer, may influence metastatic behavior. Second, we developed a rheological model and a Monte Carlo simulation framework that formalize the relationship between inflammation, blood viscosity, and metastatic competence, allowing us to test whether PAD-associated vascular remodeling can generate a threshold-based regime that suppresses metastatic dissemination.

### 1.1. Work contributions

This study offers two main contributions. First, we present clinical evidence that cancers, particularly lung cancers, arising in the setting of peripheral arterial disease and aneurysmal vasculopathy exhibit an unexpectedly low metastatic burden, indicating that vascular remodeling may directly modulate metastatic behaviour. Second, we propose and computationally evaluate a phenomenological biophysical model that links inflammation-driven alterations in blood rheology to metastatic suppression, demonstrating through Monte Carlo simulations that PAD-associated hyperviscosity can exceed a critical threshold that mechanically constrains metastatic dissemination.

## 2. Methods

To evaluate the real-life scenario of distant cancer spreading within a smoking-related pathologic vasculature, we developed an integrated approach based on two complementary components: (i) the retrospective analysis of a clinical patient dataset, and (ii) the formulation of a functional relationship linking key fluid-dynamic variables to metastatic competence.

### 2.1. Patient Identification and Selection

To investigate metastatic patterns in the context of peripheral arterial disease (PAD), we retrospectively examined a consecutive series of 976 patients diagnosed with both cancer and arteriopathy between January 2018 and December 2024 at the IRCCS San Matteo Hospital Foundation. From this population, we selected a cohort of 120 patients affected by cancer in association with aneurysmal disease. Comprehensive demographic and clinical data are provided in Supplementary Materials. Informed consent was routinely obtained from all patients at the time of hospital admission in accordance with institutional procedures. Clinical information was retrieved through review of operative registries, pneumological and vascular surgery reports, and hospital discharge summaries.

### 2.2. Statistical analysis

Statistical analyses were performed using the Excel add-in package (Microsoft Corp., Redmond, WA). Continuous variables were expressed as mean values ± standard deviation (SD) and compared using the Student’s *t*-test for independent samples. Nominal variables were compared using the χ^2^ test. A *p*-value < 0.05 was considered statistically significant. For all analyses, 95% confidence intervals (CIs) were computed for the cohort under study. It is important to note that this investigation was not designed as a case–control study; rather, it represents an analysis of consecutively collected retrospective data. In this framework, CIs and *p*-values provide complementary information. A CI offers a range of values within which the true population parameter is likely to lie, conveying both the magnitude and the precision of the estimated effect. Conversely, a *p*-value quantifies the probability of observing the sample data under the assumption that the null hypothesis (e.g., no difference between groups) is true. A small *p*-value (< 0.05) supports rejection of the null hypothesis, but it does not provide information on effect size or its clinical relevance. Thus, in the present retrospective setting, CIs may offer more informative insight into the variability and robustness of the findings than *p*-values alone. To further explore predictors of outcome, the entire dataset was analyzed using the JMP Partition algorithm (JMP Statistical Discovery Software, SAS Institute; https://www.jmp.com). This method identifies optimal data subdivisions by evaluating all possible splits of predictor variables, thereby revealing the most informative determinants of response distribution and event probability.

### 2.3. Biophysical modeling

Despite the biological uniqueness that orchestrates the movement of metastatic cells through the circulatory system, the transport of circulating tumor cells (CTCs) remains governed by the same fluid-dynamic principles that regulate blood flow in health and disease [13, 14]. Accordingly, a growing body of work has emphasized that metastasis is not solely a biological process, but also a biomechanical one, shaped by shear forces, vessel geometry, and the rheological properties of blood [15, 16]. Blood consists of plasma and cellular components: from a rheological standpoint, plasma behaves as a Newtonian fluid, whereas whole blood exhibits a distinctly non-Newtonian profile [17]. These deviations from Newtonian behaviour become particularly relevant at low flow velocities, within recirculation zones, and in secondary flows, conditions that are frequently encountered in structurally abnormal or diseased vessels, where viscosity-dependent transport and disturbed hemodynamics critically influence cellular arrest, survival, and extravasation [14, 17]. As a consequence, blood flow dynamics in pathological settings cannot be fully captured by the classical Navier–Stokes equations for linear viscous fluids.

In this study, we therefore sought to integrate established principles of hemodynamics with the dual pathological context of vasculopathy, particularly arterial wall disease such as aneurysmal degeneration, and cancer progression. Our aim was to develop a formal biophysical framework capable of recapitulating realistic disease onset and metastatic behaviour by explicitly linking vascular remodeling, blood rheology, and CTC dynamics, thereby complementing biological descriptions of metastatic dissemination with a physics-based perspective. To formalize this relationship, we developed a biophysical model that incorporates the principal hemodynamic determinants acting on CTCs. PAD and aneurysmal arteriopathy profoundly alter blood flow through modifications in vessel geometry, apparent viscosity, wall compliance, and local inflammatory burden, all of which influence CTC survival, arrest, and extravasation. In the microcirculation, blood behaves as a non-Newtonian fluid whose apparent viscosity depends on shear rate, vessel diameter, temperature, hydration state, and the extent of intravascular inflammation. We express this dependence as:

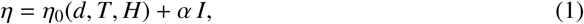

where η_0_ represents the baseline viscosity determined by physical parameters, *I* denotes the degree of vascular inflammation, and α quantifies the amplification of viscosity induced by inflammatory mediators and aggregated cellular elements such as neutrophils, platelets, and NETs. The parameter α*_I_* was introduced as a phenomenological coefficient describing the strength of coupling between inflammatory burden and apparent blood viscosity. Distinct values were assigned to control and PAD conditions to reflect the enhanced rheological sensitivity observed in chronically inflamed and structurally remodeled vasculature. These values were not calibrated to patient-specific measurements but were selected to represent relative differences between vascular contexts and to explore regime-dependent behavior. To model metastatic competence, we define a function *M* linking cancer invasiveness to these rheological conditions:

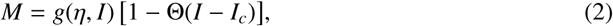

where *g*(η, *I*) describes the contribution of viscosity and inflammation to CTC survival and endothelial interactions under physiological or moderately perturbed flow. The Heaviside term Θ(*I* − *I_c_*) introduces a critical inflammatory threshold *I_c_*. When vascular inflammation and structural remodeling exceed this threshold, as is characteristic of advanced PAD or aneurysmal disease, the vasculature transitions from a permissive conduit to a mechanical and rheological filter. Under such conditions, the probability of successful CTC extravasation is strongly reduced in the model, despite elevated systemic inflammation. Below this threshold (*I* < *I_c_*), blood rheology remains compatible with CTC circulation, adhesion, and extravasation, thereby supporting metastatic dissemination. Conversely, once the threshold is crossed (*I* ≥ *I_c_*), pronounced increases in viscosity, disrupted flow patterns, endothelial stiffening, and the formation of intraluminal NET–thrombus complexes impede CTC transit and barrier crossing, effectively suppressing metastasis. This biophysical model synthesizes and explains our clinical observations, wherein cancers arising in patients with marked PAD or aneurysmal disease exhibit unexpectedly low metastatic rates. It provides a mechanistically grounded framework illustrating how pathological vascular remodeling reshapes the biomechanical environment through which CTCs must travel, thereby constraining cancer invasiveness (see Appendix Appendix A.1 for implementation details).

## 3. Results

### 3.1. Population Characteristics and Clinical Analysis

The median age of selected patient cohort at cancer diagnosis was 72.88 years. Twenty patients were female, whereas the majority were male (83.3%). Most individuals were active or former smokers. Only 18 patients (15%) were never-smokers. Among these, 11 were women (61.11%), predominantly diagnosed with breast cancer (7 cases) or gynecological malignancies (3 cases). Two patients presented with dual primary tumors: one with lung and prostate cancer, and one with breast and endometrial cancer (Fig. 1A–B). We categorized cases into three major groups: lung cancer (LC; 39 patients, 32.5%), other solid tumors with distinct primary sites (66 patients, 55%), and hematologic malignancies (15 patients, 12.5%). The majority exhibited inflammatory intra-abdominal aortic aneurysm (iAAA), although in some patients aneurysmal disease affected multiple arterial districts. Endovascular repair was performed in 43.8% of cases, and in all instances occurred after the cancer diagnosis. With respect to LC, the vast majority (36 patients) had non-small cell lung cancer (NSCLC), whereas three cases were small cell lung cancer (SCLC). Among NSCLC tumors, 23 (63.88%) were adenocarcinomas (ADC) and 14 (36.11%) were squamous cell carcinomas (SCC). All LC patients were current or former smokers, and aneurysmal disease had been documented prior to LC onset. Remarkably, across all epithelial solid tumors, only 14 cases (13.3%) exhibited metastatic disease at diagnosis, and merely 4 patients (3.8%) presented with more than two metastatic sites (Fig. 1C). Among LC cases, six patients were classified as stage IV disease. However, only three (7.6%) exhibited extrathoracic metastatic dissemination, comprising two cases of small-cell lung cancer with bone involvement and one case of non-small-cell lung cancer with hepatic metastasis. Such a rate was markedly lower than the expected prevalence of approximately 30% reported in unselected lung cancer populations. Intrathoracic metastatic patterns classified as M1a (including pleural or pericardial involvement and contralateral pulmonary nodules) were evaluated separately; their inclusion in the metastatic endpoint did not alter the direction or statistical significance of the observed association between arteriopathy and reduced metastatic burden. PD-L1 tumor proportion score (TPS) was available for 20 out of the 36 NSCLC patients. High PD-L1 expression (TPS ≥ 50%) was observed in 10 cases, intermediate expression (5% ≤ TPS < 50%) in 5 cases, and low expression (TPS = 1%) in 5 cases. The median PD-L1 expression level was 30.03% (Fig. 1D).

**Figure 1:**
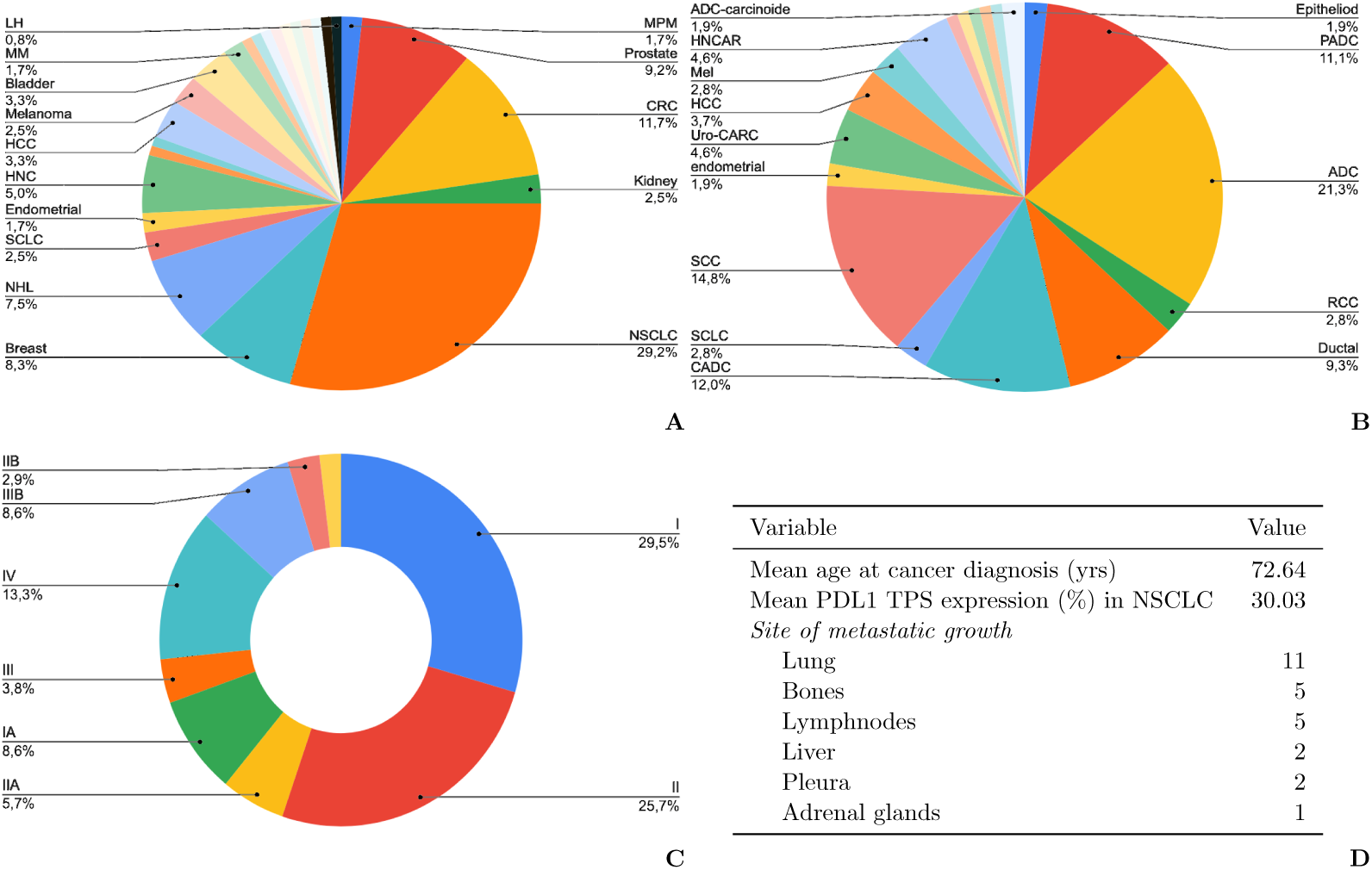
Population enrolled. A-B) Primary site of cancer origin, histotype; C) disease stage (TNM 8 ed) of the cases evaluated; D)data on metastatic sites and PDL1expression referred to NSCLC. MM: multiple myeloma, NHL: non-Hodgking Lymphoma, CRC: colorectal cancer, MPM: malignant pleural mesothelioma, HNCAR: head and neck carcinoma, uro-CARC:urothelial carcinoma; PADC: pancreatic adenocarcinoma; RCC: renal cell carcinoma; Ductal: ductal breast cancer; CADC: cervical adenocarcinoma; mel: melanoma.

### 3.2. Patient Biochemical Profile

We then examined the biochemical profile of each case, focusing on variables most closely linked to inflammation and atherosclerosis. For all enrolled patients, values for platelet count (PLT) and serum total cholesterol were available, with mean levels of 216.4 × 10^9^/L (PLT) and 141 mg/dL (total cholesterol). The mean HDL cholesterol level was 46.33 mg/dL, and no significant changes were observed between baseline and the time of cancer diagnosis. In a subset of 25 patients, LDL cholesterol values were available, with a mean of 74.64 mg/dL. Overall, serum C-reactive protein (CRP), a systemic marker of inflammation, was unexpectedly low at cancer diagnosis (mean value: 1.22 mg/dL), suggesting that the inflammatory process associated with advanced vasculopathy may be predominantly localized to the vascular wall rather than reflected by systemic biomarkers. Diabetes mellitus was reported in 27 patients (22.5%). Regarding respiratory function, a diagnosis of COPD was established in 41 of the 120 cases, whereas in the remaining 79 patients spirometry was not performed despite a frequently long-standing smoking history (Fig. 2). These findings were then compared with those from an unselected lung cancer population (male: 70.69%) diagnosed and followed in our Institution over the same time interval. The vast majority of tumors were adenocarcinomas (60.65%), and the prevalence of metastatic disease at diagnosis was 48.27%. In this population, the primary tumor site was the lung in most cases (NSCLC 75.48%, SCLC 5.74%), whereas in about 10% of patients lung lesions represented secondary metastases from other organs (predominantly breast cancer, 4%).

**Figure 2:**
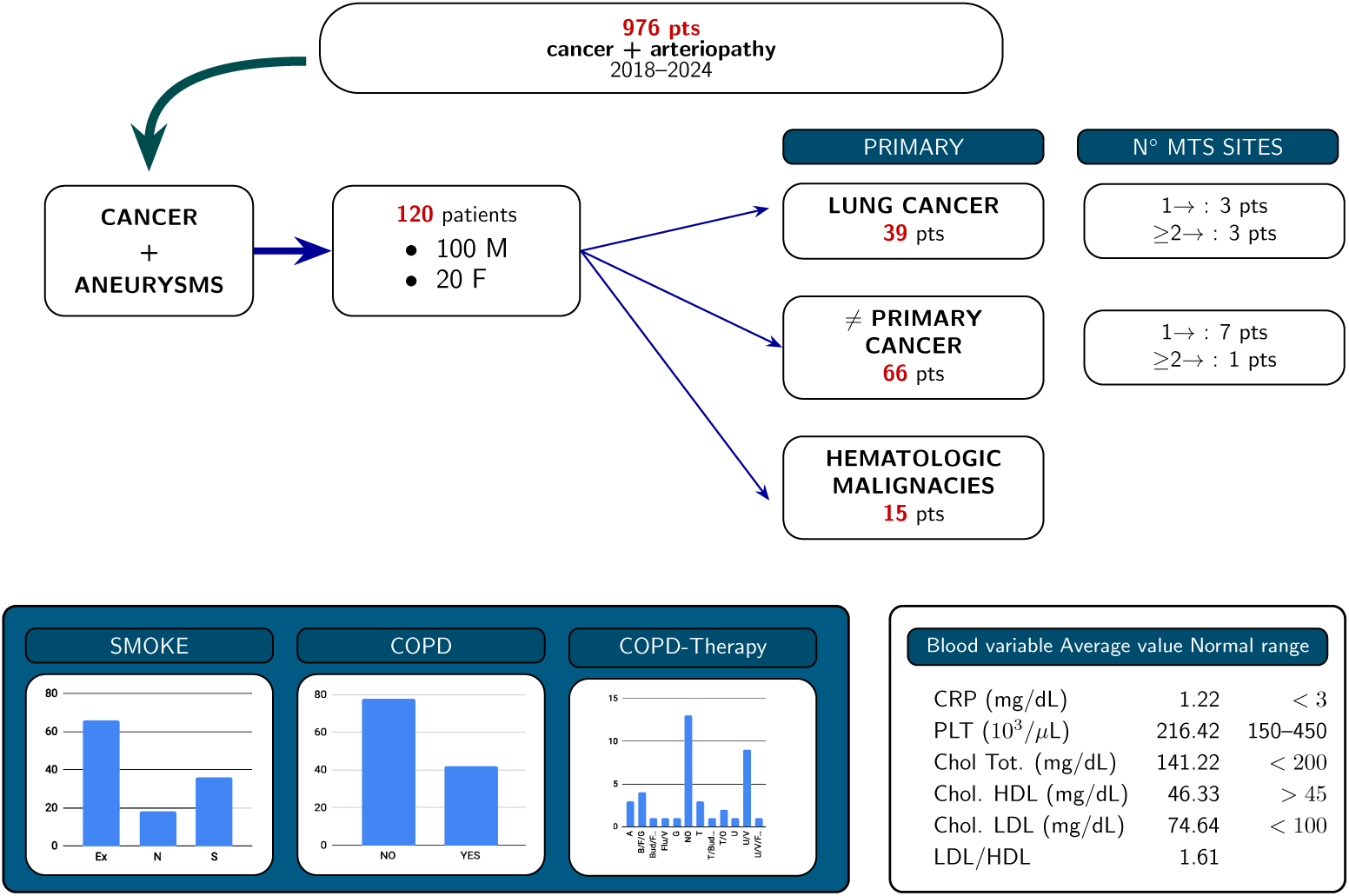
Study design and demographic, clinical and biochemical features of the population evaluated.

The null hypothesis of no difference in metastatic rates between (i) the study cohort and (ii) the unselected lung cancer cohort followed at the Respiratory Diseases Unit was rejected, with a chi-square statistic of χ^2^ = 10.37 and a corresponding *p* value of 0.0012, indicating statistical significance at the conventional threshold (*p* < 0.05). The finding demonstrated that the reduced metastatic burden observed in the study cohort differed significantly from the rate expected in an unselected lung cancer population. In contrast, no statistically significant difference was detected between the institutional reference lung cancer cohort (Yates-corrected χ^2^ = 3.714, *p* = 0.5396) and metastatic rates reported in the literature [18, 4], indicating consistency between the reference cohort and published data. Consistently, when the study cohort was directly compared with literature-derived metastatic rates, the difference remained statistically significant (χ^2^ = 5.5588, *p* = 0.018), further supporting the robustness of the observed reduction in metastatic dissemination associated with the study population.

To further strengthen these findings, we retrospectively evaluated the incidence of cancer among unselected patients hospitalized in the Vascular Surgery Unit between January 1*^st^* 2024 and January 31*^st^* 2025. A total of 171 patients were assessed, of whom 15.78% were female. Cancer was diagnosed in 21.63% of cases, with the majority (59.45%) detected at an early stage. Lung cancer accounted for approximately 30% of diagnoses in this cohort, and metastatic dissemination was present in about 50% of cases. Although the number of cases was limited, this metastatic proportion is consistent with that reported in the literature for unselected lung cancer population. When this Vascular Surgery cancer cohort was compared with the study cohort, the null hypothesis of no difference in metastatic rate was again rejected (χ^2^ = 17.15, *p* = 3.5 × 10^−5^), indicating a significantly lower metastatic burden in the study population. In contrast, no statistically significant difference was observed between the Vascular Surgery cancer cohort and literature-reported metastatic rates (χ^2^ = 1.40, *p* = 0.23), supporting the representativeness of this cohort as an internal control. The 95% confidence intervals for metastatic rates further supported this interpretation: 5.9–30.5 for the study cohort, 13.9–28.7 for the unselected lung cancer population followed in the Respiratory Disease Unit, and 15.7–28.6 for unselected cancer patients diagnosed in the Vascular Surgery Unit. Collectively, these comparisons indicate that the reduced metastatic burden is specific to the study cohort and not attributable to institutional, diagnostic, or selection bias.

### 3.3. Partition Analysis

To further validate our findings, we performed a predictor analysis to identify the most relevant variables associated with tumor evolution and to explore potential splits linked to metastatic burden, using the resulting decision trees to highlight markers of translational interest. In the first model, the dataset was analyzed without any preselection for arteriopathy, gender, or smoking status. In this context, the age at cancer diagnosis (younger than 67 years) and total cholesterol level (threshold 175 mg/dL), followed, as expected, by histotype, emerged as the main factors associated with tumor progression, measured as the number of distant metastases (Fig. 3). When arterial disease was introduced as a covariate, it became the dominant predictor (Fig. 4), followed by intrinsic tumor biology (histotype), biochemical profile, and smoking habit.

**Figure 3:**
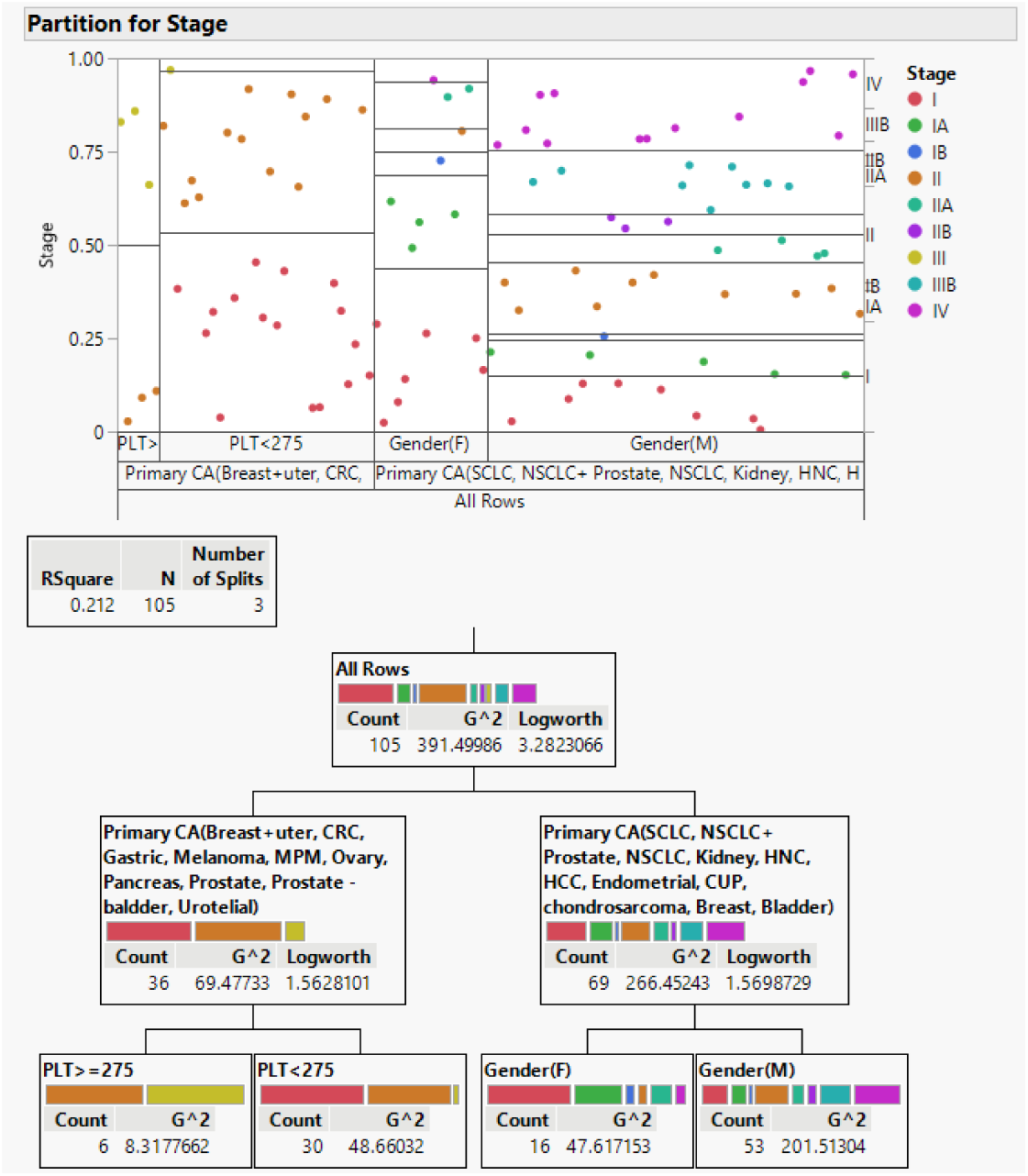
Partition analysis (excluding arteriopathy) for disease stage: most relevant splits regard tumor primary site, PLT and gender.

**Figure 4:**
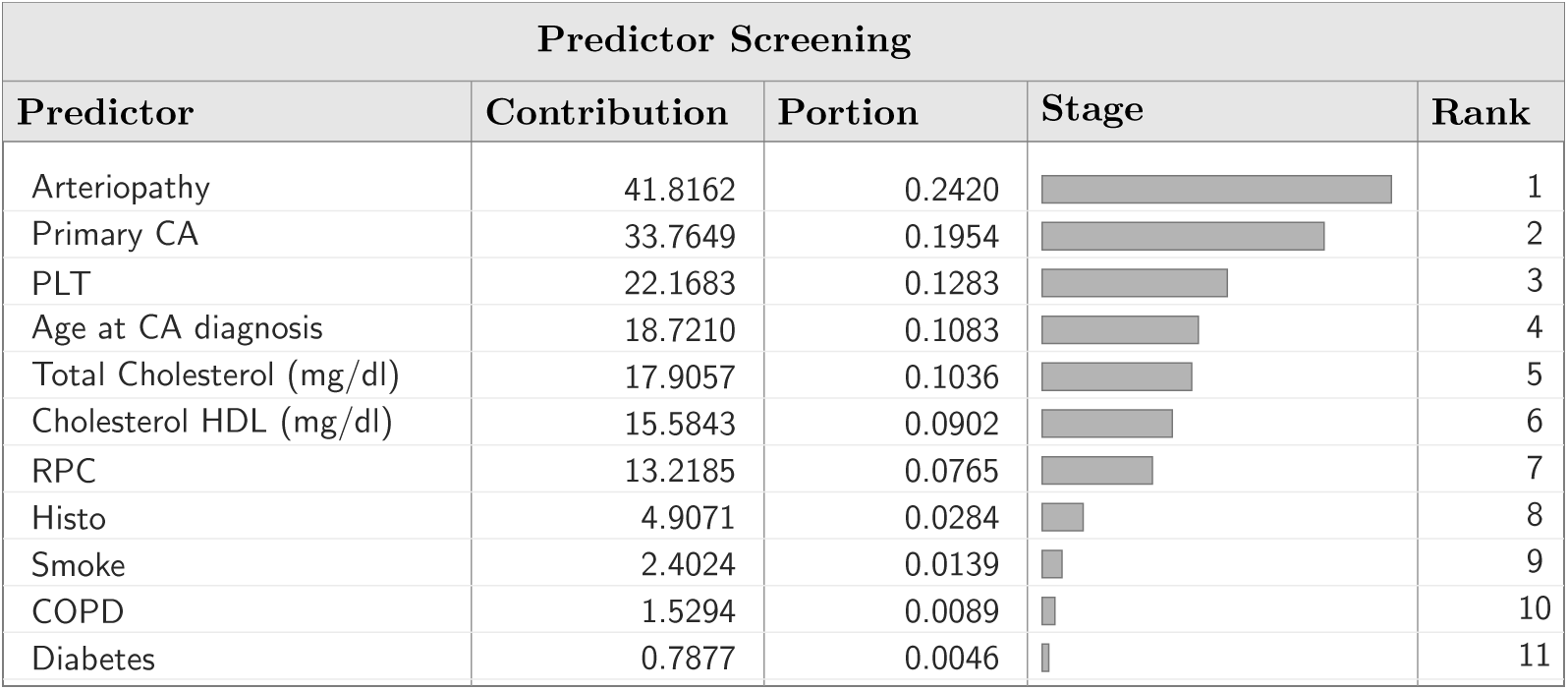
Partition analysis for disease stage: the screening underlines that arteriopathy is the most relevant variable associated to outcome (number of metastases/stage).

### 3.4. Biophysical Model Results

In the Monte Carlo test, each virtual patient was assigned an inflammatory value *I* sampled from a truncated normal distribution characteristic of the vascular context:

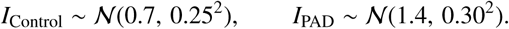

For each sampled value of inflammation, viscosity was computed using

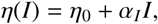

with weaker rheological coupling in the Control condition (α*_I_* = 0.7) and stronger coupling in PAD (α*_I_* = 1.4). The metastatic probability was then evaluated as

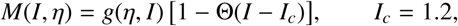

where the sub-threshold term was defined as

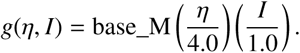

where base_M is the baseline metastatic probability scale that applies only when inflammation is below the critical threshold *I_c_*. For *I* ≥ *I_c_*, the Heaviside function Θ set *M* = 0, suppressing metastasis entirely. A Bernoulli trial with probability *M*(*I*, η) determined metastatic outcome for each simulated patient. Repetition of this process across many Monte Carlo runs generated distributions of metastatic rates for Control and PAD conditions, allowing quantitative comparison of metastatic burden under the two rheological regimes. 20,000 virtual patients for the Control cohort, and 20,000 virtual patients for the PAD cohort, were simulated. The simulation confirmed a substantial reduction in predicted metastatic burden under PAD conditions compared with the control vascular environment (Fig. 5). In the control scenario (cancer without PAD), the mean metastasis rate was 0.169, with a narrow standard deviation (0.003) and a 95% confidence interval of 0.165–0.175, indicating a stable and reproducible metastatic output (Table 1).

**Figure 5:**
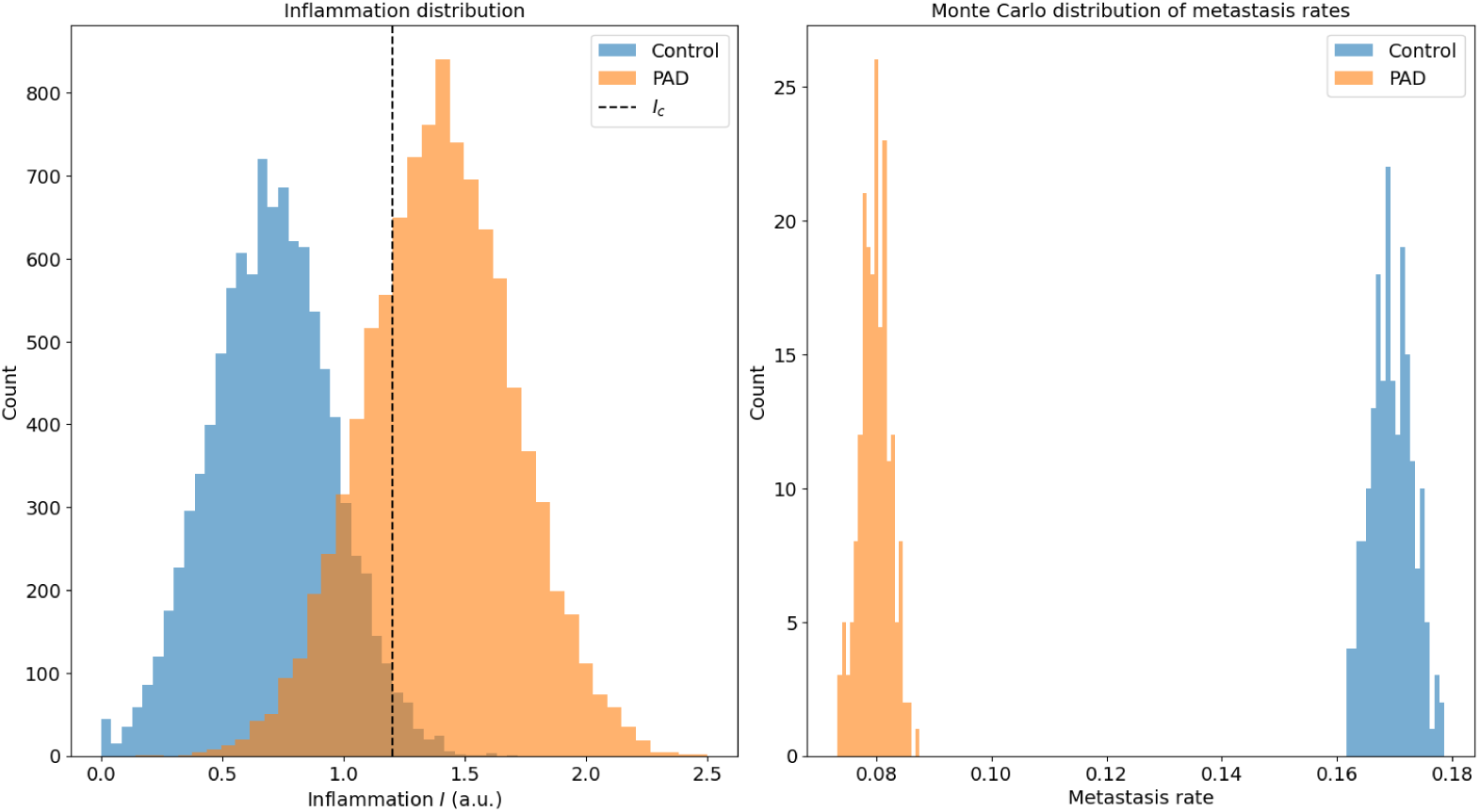
Simulation results. Inflammation is expressed in arbitrary units (a.u.). *I_c_* is the inflammation threshold.

**Table 1:** Monte Carlo simulation results comparing metastatic rates in Control vs. PAD conditions.

| Condition | Mean metastasis rate | SD | 95% CI |
| --- | --- | --- | --- |
| Control (cancer without PAD) | 0.169 | 0.003 | [0.165, 0.175] |
| PAD (cancer + aneurysm) | 0.080 | 0.002 | [0.077, 0.083] |

In contrast, under PAD-like rheological conditions, the mean metastasis rate decreased to 0.080, with similarly tight variability (SD = 0.002; 95% CI: 0.077–0.083). These findings demonstrate that, across repeated stochastic realizations, PAD consistently and robustly suppresses metastatic dissemination by approximately 50%, in agreement with the threshold-dependent rheological mechanism encoded in the model.

A sensitivity analysis was performed by varying the inflammation–viscosity coupling parameter α*_I_*for PAD-like conditions from 1.0 to 2.0 in steps of 0.05, while keeping all control parameters fixed. Across the entire range of tested values, PAD-like simulations consistently yielded lower metastatic probabilities than the control scenario (Fig. 6). According ti the previous test, at the baseline setting α*_I_* = 1.40, the model predicted a mean metastatic rate of 0.080 in PAD-like conditions compared with 0.169 in controls, corresponding to a relative risk of 0.47 and a 52.6% reduction in metastatic probability. Notably, the mean metastatic rate in PAD-like simulations remained lower than that of controls for 100% of tested α*_I_*values, with complete non-overlap of 95% confidence intervals across the full parameter range. Although the predicted metastatic rate showed a modest positive trend with increasing α*_I_*(slope *b* = 0.0167 per unit α*_I_*), the overall suppression of metastasis in PAD-like conditions was preserved, indicating that the observed threshold-dependent effect does not rely on fine-tuning of model parameters.

**Figure 6:**
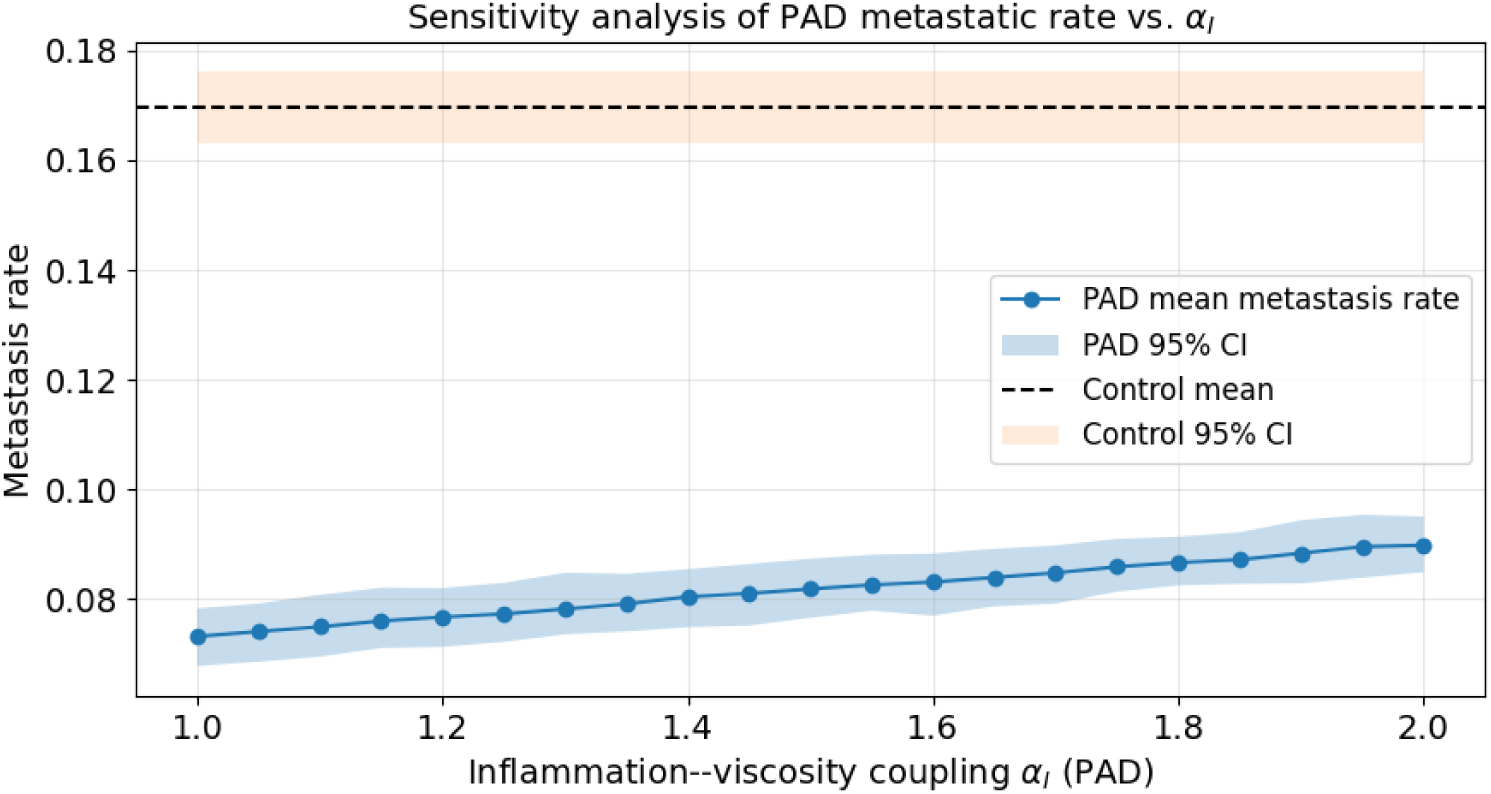
Simulation results. Sensitivity analysis of α*_I_* PAD varying from 1.0 to 2.0 (steps: 0.05). At each step, Montecarlo simulation over 10000 cases was run (PAD 95% CI.

As a robustness check, we replaced the idealized Heaviside switch in the metastatic competence function with a smooth logistic transition centered at the same critical inflammatory threshold *I_c_* (cfr. eq. 2). In this alternative formulation, metastatic probability decreased progressively as inflammation increased beyond *I_c_*, rather than being abruptly set to zero. Monte Carlo simulations under this smooth-transition regime yielded results that were quantitatively comparable to those obtained with the Heaviside formulation. Specifically, the mean predicted metastatic rate in PAD-like conditions was 0.089 (95% CI: 0.084–0.094), compared with 0.166 (95% CI: 0.160–0.173) in the control scenario, corresponding to a relative reduction of approximately 46%. Importantly, confidence intervals remained non-overlapping between PAD-like and control simulations. These findings indicate that the threshold-dependent suppression of metastatic probability observed in PAD-like conditions reflects a robust regime shift in the model rather than an artifact of the specific mathematical form of the transition function.

## 4. Discussion

Cancer dissemination is classically framed as the convergence of oncogenic drivers, permissive microenvironments, and systemic inflammatory cues, with lung cancer (LC) representing a paradigmatic model of aggressive metastatic behavior [1, 3, 4]. Most contemporary work has focused on tumor-intrinsic drivers [19, 20, 21, 10, 18, 22], as well as on the molecular organization of the pre-metastatic niche [8, 23]. In parallel, an emerging body of literature has begun to emphasize the contribution of physical and rheological factors, showing that shear forces, vessel architecture, and fluid properties critically shape metastatic trajectories [15, 16, 24, 25]. Within this framework, blood is increasingly recognized not as a passive carrier but as an active, dynamic medium whose viscosity, non-Newtonian behavior and flow patterns can modulate cancer cell fate [17, 26, 27]. Our work extends this perspective into a specific clinical niche, LC and other solid tumors arising in the context of smoke-associated PAD, with aneurysmal disease, by showing that advanced vasculopathy can be associated with an unexpectedly low metastatic burden and by proposing a mechanistic, physics-based correlation function that accounts for this phenomenon.

Several epidemiological and vascular studies have documented a non-trivial relationship between LC and aneurysmal disease (Fig. 7). Patients with LC exhibit a higher prevalence of abdominal aortic aneurysms (AAA) than the general population, and vice versa, suggesting shared exposure to tobacco and overlapping inflammatory and vascular risk profiles rather than a simple coincidence [28, 29, 30, 31, 32, 33]. At the same time, detailed metastasis patterns in such populations have rarely been dissected. Our retrospective analysis indicates that, in a cohort of patients with cancer and aneurysmal PAD, the rate of distant metastases at diagnosis is significantly lower than in an unselected LC population from the same institution and also lower than expected from large series describing stage distribution and metastatic patterns in LC [2, 3, 4]. Importantly, the reduced metastatic burden persisted when both any metastatic disease (M1) and the stricter endpoint of extrathoracic dissemination (M1b/M1c) were considered, supporting the robustness of the observed association. This observation is not easily explained by differences in histotype, sex distribution, or exposure to oncogenic drivers alone. Rather, it argues that the vascular phenotype, characterized by aneurysmal dilation, chronic inflammation of the arterial wall, and altered hemodynamics, actively constrain metastatic dissemination.

**Figure 7:**
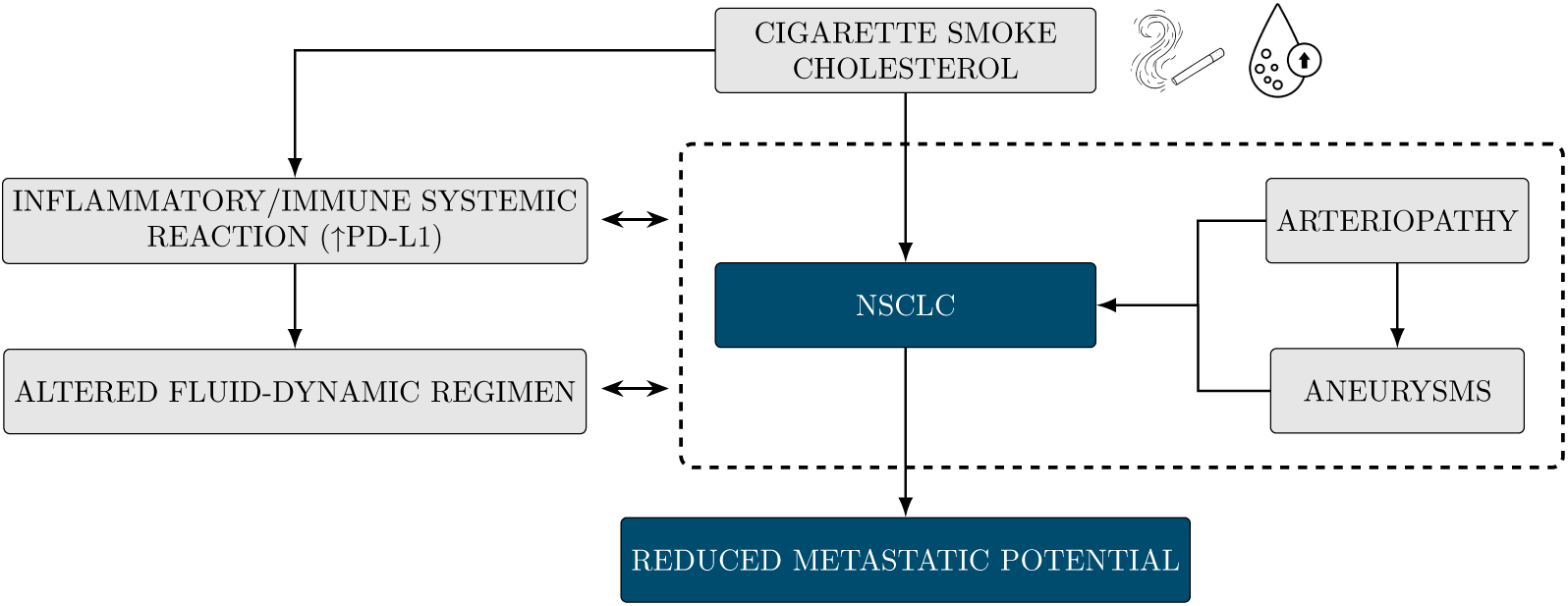
Pathogenic mechanism of NSCLC progression in case of PAD.

From a mechanistic standpoint, numerical models embed concepts that are increasingly recognized across vascular biology and hemorheology. Aneurysmal disease is associated with elastin degradation, collagen remodeling, wall inflammation and immune infiltration, collectively leading to arterial stiffening, altered compliance and complex flow patterns including vortices, re-circulation zones and low-shear regions [34, 35, 36, 14, 13]. Blood itself behaves as a non-Newtonian fluid, and its apparent viscosity depends on shear rate, vessel diameter and inflammatory state [17, 37, 38]. Classical hemorheological studies in cancer have shown that increased whole-blood viscosity and altered microrheology can correlate with worse outcomes and higher metastatic propensity [39, 40, 41, 42, 43]. More recent work has refined this view by showing that extracellular fluid viscosity can enhance cell migration and dissemination in a non-linear, context-dependent fashion [25]. Together, these data suggest that viscosity and flow are not uniformly pro-metastatic or anti-metastatic, but act through threshold-like and regime-dependent behaviours. Specifically, while moderate increases in viscosity and extracellular fluid resistance can enhance cell migration and metastatic competence under certain conditions [24], extreme or chronic rheological alterations may instead impose mechanical constraints on cellular transit. This conceptual shift is supported by recent theoretical perspectives highlighting the existence of regime-dependent or history-dependent mechanical states in biological fluids and tissues, including the notion of a “memory of viscosity” [15, 16].

In our model, we formalized blood viscosity as a function of baseline rheology and inflammation, η = η_0_ + α*_I_ I*, and introduced a critical inflammatory threshold *I_c_* such that, when exceeded, the vasculature transitions from a metastasis-permissive conduit into a rheological and mechanical filter. Below *I_c_*, moderate hyperviscosity and disturbed flow can, in principle, favour CTC arrest, adhesion and extravasation, in line with reports linking elevated viscosity and pro-thrombotic states to metastatic progression [43, 42, 44]. Above *I_c_*, however, the model predicts that extreme hyperviscosity, wall stiffening, NET–thrombus structures and local turbulence conspire to impair CTC trafficking and survival. Monte Carlo simulations, in which inflammatory states were sampled from distributions representative of control and PAD conditions, consistently reproduced this threshold effect: in the PAD-like setting, where *I* frequently exceeded *I_c_* and the coupling between inflammation and viscosity (α*_I_*) was stronger, the predicted metastatic rate was approximately halved compared to the control scenario (mean 0.080 vs. 0.169, with narrow confidence intervals), despite identical intrinsic pro-metastatic scaling (base_M) between the two contexts. Importantly, sensitivity analysis demonstrated that the PAD-associated suppression of metastatic probability was robust across a broad range of inflammation–viscosity coupling strengths, supporting the interpretation of a regime-dependent rheological effect rather than a parameter-specific artifact. This quantitative result supports the notion that it is the vascular and rheological regime, rather than tumor biology alone, that drives the observed attenuation in metastatic burden.

The biochemical and immunological profile of our cohort further supports a locally organized, vasculopathy-driven rheological state rather than a diffuse systemic inflammatory syndrome. While platelet counts and conventional systemic markers such as CRP and lipid fractions (total cholesterol, HDL, LDL, LDL/HDL ratio) did not show marked elevations, PD-L1 expression in tumors was frequently high, and the literature indicates that PD-1/PD-L1 signalling is intricately involved not only in cancer immune escape but also in vascular inflammation, aneurysm progression and neointimal hyperplasia [45, 46, 47]. Moreover, the complex interplay between smoking, vascular inflammation, atherosclerosis, AAA risk and LC development is well documented [48, 49, 50, 51, 33]. In this context, our findings suggest that, in a subset of smokers with advanced PAD, the vasculature may reach a pathologic but metastasis-restrictive state in which structural and rheological derangements supersede the pro-metastatic advantages of moderate hyperviscosity.

The role of blood rheology in cancer progression has long been recognized [41]. Yet it has traditionally been interpreted within a largely monotonic framework in which increased viscosity and altered fluid-dynamics properties are assumed to uniformly promote tumor dissemination. Early clinical studies demonstrated that cancer patients frequently exhibit elevated whole-blood viscosity, increased erythrocyte aggregation, and reduced red blood cell deformability, and these alterations were associated with advanced disease stage and poor prognosis [42]. Rheological abnormalities have been primarily viewed as facilitators of metastatic spread through enhanced tumor cell arrest and microvascular obstruction. More recent work has reinforced the physical dimension of metastasis, framing circulating tumor cell (CTC) transport, arrest, and extravasation as flow-dependent processes governed by shear forces, vessel geometry, and non-Newtonian blood behavior [15, 16]. Computational and experimental studies have further shown that blood viscosity interacts nonlinearly with vascular architecture, generating recirculation zones, low-shear regions, and heterogeneous flow patterns that critically influence cellular transport [38, 26, 17]. In parallel, studies on cancer-associated immunothrombosis have revealed that advanced inflammatory states characterized by platelet activation, fibrin deposition, and neutrophil extracellular trap formation can simultaneously promote and restrict tumor dissemination, depending on spatial and temporal context [44].

The study presents some limitations. The clinical analysis was retrospective and relied on routinely collected imaging and laboratory data rather than on prospectively acquired vascular or rheological measurements. Consequently, the mechanistic interpretation proposed here cannot yet be directly validated against in vivo assessments of blood viscosity, microcirculatory flow characteristics, endothelial permeability, or aneurysmal wall biomechanics in the same patients. Moreover, the physics-based model and the Monte Carlo simulations were conceived as a phenomenological framework grounded in established principles of rheology and vascular biology, but they were not calibrated or validated using experimental measurements or patient-specific biophysical data. Importantly, the numerical values assigned to α*_I_* do not represent direct physiological measurements, but rather encode relative differences in inflammation–rheology coupling between control and PAD-like vascular states. The qualitative behavior of the model was robust across a broad range of α*_I_* values, indicating that the observed threshold effect does not rely on fine-tuning. Thus, the model predictive capacity should be regarded as hypothesis-generating rather than confirmatory. Inflammation in the model was represented as a single scalar variable, whereas real vascular inflammation encompasses a multidimensional set of immune, stromal, and extracellular matrix alterations that are only partially captured in a reduced formulation. Finally, the cohort size was relatively limited and demographically skewed due to the epidemiology of smoking-related PAD, potentially restricting the generalizability of the findings and concealing sex- or age-specific vascular effects. In addition, incomplete pulmonary function assessment, including the absence of spirometry in many long-term smokers, prevented a detailed evaluation of the interplay between COPD, inhaled therapies, bronchial inflammation, and vascular rheology. Overall, while the convergence between clinical observations and model predictions is notable, future prospective studies incorporating direct rheological, hemodynamic, and structural measurements will be essential to validate the threshold-based mechanism proposed here.

## 5. Conclusions

By integrating retrospective clinical evidence with a biophysical modeling approach, this study suggests that chronic PAD and aneurysmal vasculopathy may reshape the circulatory microenvironment in a manner that constrains metastatic dissemination across solid tumors. The unexpectedly low metastatic burden observed in patients with advanced vasculopathy supports the hypothesis that, beyond a critical threshold, inflammation-driven alterations in blood rheology and vascular biomechanics can shift circulation from a metastasis-permissive conduit to a metastasis-restrictive mechanical filter. While the phenomenological model and Monte Carlo simulations presented here have not yet been extensively validated against direct rheological or hemodynamic measurements, they provide a coherent mechanistic interpretation of the clinical findings and, importantly, define a prospective and testable framework for future investigation. By formalizing a threshold-based, rheology-driven mechanism of metastatic restraint, this work motivates prospective studies combining direct hemorheological assessment, high-resolution vascular imaging, and longitudinal evaluation of circulating tumor cell dynamics to determine whether vasculopathy- and rheology-informed parameters may ultimately contribute to metastatic risk stratification and clinical decision-making.

## Data Availability

Data will be available if properly required to the PI

https://docs.google.com/spreadsheets/d/1dPA8UhURGtbF8tzRz01c4usK6LBqiWEfVzDiUq4D4Vs/edit?usp=share_link

## Author Contributions

Conceptualization, GMS, CN, FRB, VA, AB; Methodology, GMS, CN, FRB, VC, IF, IL, VA, AB; Software, GMS; Validation, GMS, FRB, AGC, AB.; Formal Analysis and investigation, GMS, CN, FRB, IF, VC, TG, IS, VA; Data Curation, GMS, CN, FRB, VA, AB; Writing – Original Draft Preparation, GMS, CN, FRB; Writing – Review & Editing, GMS, FRB, AGC, AB PC;, Funding Acquisition, GMS.

## Informed Consent Statement

This is not a clinical trial. Informed consent from each patient was routinely collected at hospital admission in accordance with standard hospital procedures.

## Data Availability Statement

The datasets presented in this study can be found in online repositories. The names of the repository/repositories and accession number(s) can be found in the article/supplementary material.

## Appendix A. Appendix

### Appendix A.1. Numerical Implementation of the Biophysical Model

The phenomenological model linking PAD-related rheology to metastatic dissemination was implemented in Python. The numerical framework translated the model *M*(η, *I*) into a patient-level stochastic simulator, allowing comparison between different vascular contexts (control vs. PAD). Inflammation was represented by a scalar variable *I*, and the transition between metastasis-permissive and metastasis-suppressive regimes was encoded through the Heaviside step function Θ(*x*), defined as:

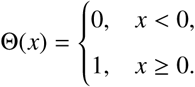

This function was used to enforce the critical inflammatory threshold *I_c_* in the correlation function

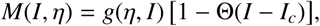

as detailed in the previous section. Each vascular context was characterized by a set of parameters grouped in a `ContextParams` data structure, including: the inflammatory threshold *I_c_*, the baseline viscosity η_0_, the coupling coefficient α*_I_* between inflammation and viscosity, the baseline metastatic propensity (`base_M`) in the sub-threshold regime, and the mean and standard deviation of the inflammatory state (*I*_mean_, *I*_sd_) in the population. Within a given context, the apparent viscosity was modeled as:

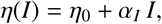

in accordance with the simplified rheological relation introduced earlier. The baseline viscosity parameter η_0_ was set to 3.5 in normalized units, corresponding to a physiologically plausible apparent whole-blood viscosity under low-to-moderate inflammatory conditions. This parameter serves as a reference state, upon which inflammation-driven rheological amplification is super-imposed, rather than as a direct measurement in physical units.

The metastatic dissemination probability for an individual virtual patient was given by

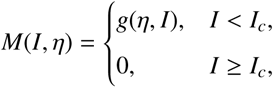

where *g*(η, *I*) was implemented phenomenologically as

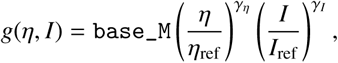

with η_ref_ and *I*_ref_ used for normalization. The resulting value *M*(*I*, η) was interpreted as a Bernoulli probability. Population-level behaviour was obtained by Monte Carlo sampling. For each context, a cohort of *n* virtual patients was generated by drawing inflammatory states *I* from a truncated normal distribution

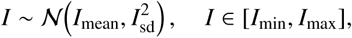

where *I*_min_ and *I*_max_ enforced physiologically plausible bounds. For each sampled value of *I*, the model computed η(*I*) and the corresponding metastatic probability *M*(*I*, η), and then generated a binary outcome (metastasis: yes/no) from a Bernoulli distribution with parameter *M*(*I*, η). The proportion of metastatic outcomes in the simulated cohort provided an estimate of the metastatic rate for that vascular context. Two archetypal scenarios were examined: (i) a *control* condition, representing cancers without PAD and characterized by lower mean inflammation and weaker coupling between *I* and η; and (ii) a *PAD* condition, representing cancers in patients with aneurysmal disease, with higher mean inflammation and stronger inflammation–viscosity coupling (hyperviscosity). Both scenarios shared the same intrinsic metastatic scale (`base_M`), so that differences in simulated metastatic burden arose from the distribution of *I* and the activation of the threshold *I_c_*. For each scenario, the code reported summary statistics for *I*, η, and the metastatic rate, and optionally generated illustrative plots of inflammation distributions, *M*(*I*) curves, and predicted metastatic fractions. This numerical implementation therefore provided a concrete, testable realization of the proposed physics-based correlation function linking vascular pathology, blood rheology, and metastatic behaviour.

## References

[1] S. Xie, Z. Wu, Y. Qi, B. Wu, X. Zhu, The metastasizing mechanisms of lung cancer: Recent advances and therapeutic challenges, Biomedicine & Pharmacotherapy 138 (2021) 111450. doi:10.1016/j.biopha.2021.111450.

[2] R. L. Siegel, A. N. Giaquinto, A. Jemal, Cancer statistics, 2024, CA: A Cancer Journal for Clinicians 74 (2024) 12–49. doi:10.3322/caac.21820.

[3] A. A. Thai, B. J. Solomon, L. V. Sequist, J. F. Gainor, R. S. Heist, Lung cancer, The Lancet 398 (10299) (2021) 535–554. doi:10.1016/S0140-6736(21)00312-3.

[4] T. Xie, B. M. Qiu, J. Luo, Y. F. Diao, L. W. Hu, X. L. Liu, Y. Shen, Distant metastasis patterns among lung cancer subtypes and impact of primary tumor resection on survival in metastatic lung cancer using seer database, Scientific Reports 14 (1) (2024) 22445. doi:10.1038/s41598-024-73389-6.

[5] F. Fu, Z. Chen, H. Chen, Treating lung cancer: defining surgical curative time window, Cell Research 33 (9) (2023) 649–650. doi:10.1038/s41422-023-00852-w.

[6] A. S. La’ah, S. H. Chiou, Cutting-edge therapies for lung cancer, Cells 13 (5) (2024) 436. doi:10.3390/cells13050436.

[7] S. D. Lin, C. Y. Tong, D. D. Huang, A. Rossi, H. Adachi, M. Miao, W. X. Zheng, J. Guo, The time-to-surgery interval and its effect on pathological response after neoadjuvant chemoimmunotherapy in non-small cell lung cancer: a retrospective cohort study, Translational Lung Cancer Research 13 (10) (2024) 2761–2772. doi:10.21037/tlcr-24-781.

[8] G. M. Stella, S. Kolling, S. Benvenuti, C. Bortolotto, Lung-seeking metastases, Cancers 11 (7) (2019) 1010. doi:10.3390/cancers11071010.

[9] A. Lundin, B. Driscoll, Lung cancer stem cells: progress and prospects, Cancer Letters 338 (1) (2013) 89–93. doi:10.1016/j.canlet.2012.08.014.

[10] X. Chu, W. Tian, J. Ning, G. Xiao, Y. Zhou, Z. Wang, Z. Zhai, G. Tanzhu, J. Yang, R. Zhou, Cancer stem cells: advances in knowledge and implications for cancer therapy, Signal Transduction and Targeted Therapy 9 (1) (2024) 170. doi:10.1038/s41392-024-01851-y.

[11] Q. Zhou, L. Zu, L. Li, X. Chen, X. Chen, Y. Li, H. Liu, Z. Sun, Screening and establishment of human lung cancer cell lines with organ-specific metastasis potential, Zhongguo Fei Ai Za Zhi 17 (3) (2014) 175–182. doi:10.3779/j.issn.1009-3419.2014.03.20.

[12] X. Guo, X. Zhu, L. Zhao, X. Li, D. Cheng, K. Feng, Tumor-associated calcium signal transducer 2 regulates neovascularization of non-small-cell lung cancer via activating erk1/2 signaling pathway, Tumour Biology 39 (3) (2017) 1010428317694324. doi:10.1177/1010428317694324.

[13] M. P. D’Arienzo, L. Rarita, Dynamics of blood flows in the cardiocirculatory system, Computation 12 (10) (2024) 194. doi:10.3390/computation12100194.

[14] S. Numata, K. Itatani, K. Kanda, K. Doi, S. Yamazaki, K. Morimoto, K. Manabe, K. Ikemoto, H. Yaku, Blood flow analysis of the aortic arch using computational fluid dynamics, European Journal of Cardio-Thoracic Surgery 49 (6) (2016) 1578–1585. doi:10.1093/ejcts/ezv459.

[15] D. Wirtz, K. Konstantopoulos, P. C. Searson, The physics of cancer: the role of physical interactions and mechanical forces in metastasis, Nature Reviews Cancer 11 (7) (2011) 512–522.

[16] G. Follain, D. Herrmann, S. Harlepp, V. Hyenne, N. Osmani, S. C. Warren, et al., Fluids and their mechanics in tumour transit: shaping metastasis, Nature Reviews Cancer 20 (2) (2020) 107–124.

[17] S. Lynch, N. Nama, C. A. Figueroa, Effects of non-newtonian viscosity on arterial and venous flow and transport, Scientific Reports 12 (1) (2022) 20568. doi:10.1038/s41598-022-19867-1.

[18] C. Jiang, N. Zhang, X. Hu, H. Wang, Tumor-associated exosomes promote lung cancer metastasis through multiple mechanisms, Molecular Cancer 20 (1) (2021) 117.

[19] D. X. Nguyen, J. Massagué, Genetic determinants of cancer metastasis, Nature Reviews Genetics 8 (5) (2007) 341–352. doi:10.1038/nrg2101.

[20] S. A. Patel, P. Rodrigues, L. Wesolowski, S. Vanharanta, Genomic control of metastasis, British Journal of Cancer 124 (1) (2021) 3–12. doi:10.1038/s41416-020-01127-6.

[21] J. Fares, M. Y. Fares, H. H. Khachfe, H. A. Salhab, Y. Fares, Molecular principles of metastasis: a hallmark of cancer revisited, Signal Transduction and Targeted Therapy 5 (1) (2020) 28. doi:10.1038/s41392-020-0134-x.

[22] M. Mathieu, L. Martin-Jaular, G. Lavieu, C. Théry, Specificities of secretion and uptake of exosomes and other extracellular vesicles for cell-to-cell communication, Nature Cell Biology 21 (1) (2019) 9–17. doi:10.1038/s41556-018-0250-9.

[23] R. Lusby, E. Demirdizen, M. Inayatullah, P. Kundu, O. Maiques, Z. Zhang, M. G. Terp, V. Sanz-Moreno, V. K. Tiwari, Pan-cancer drivers of metastasis, Molecular Cancer 24 (1) (2025) 2. doi:10.1186/s12943-024-02182-w.

[24] K. Bera, A. Kiepas, I. Godet, Y. Li, P. Mehta, B. Ifemembi, C. D. Paul, A. Sen, S. A. Serra, K. Stoletov, J. Tao, G. Shatkin, S. J. Lee, Y. Zhang, A. Boen, P. Mistriotis, D. M. Gilkes, J. D. Lewis, C. M. Fan, A. P. Feinberg, M. A. Valverde, S. X. Sun, K. Konstantopoulos, Extracellular fluid viscosity enhances cell migration and cancer dissemination, Nature 611 (7935) (2022) 365–373. doi:10.1038/s41586-022-05394-6.

[25] P. Strzyz, The memory of viscosity, Nature Reviews Molecular Cell Biology 24 (1) (2023) 3. doi:10.1038/s41580-022-00563-x.

[26] Q. Q. Wang, B. H. Ping, Q. B. Xu, W. Wang, Rheological effects of blood in a nonplanar distal end-to-side anastomosis, Journal of Biomechanical Engineering 130 (5) (2008) 051009. doi:10.1115/1.2948418.

[27] M. Saldana, S. Gallegos, E. Gálvez, J. Castillo, E. Salinas-Rodríguez, E. Cerecedo-Sáenz, J. Hernández-Ávila, A. Navarra, N. Toro, The reynolds number: A journey from its origin to modern applications, Fluids 9 (12) (2024) 299. doi:10.3390/fluids9120299.

[28] A. Hohneck, T. Shchetynska-Marinova, G. Ruemenapf, M. Pancheva, R. Hofheinz, J. Boda-Heggemann, E. Sperk, P. Riffel, J. Michels, M. Borggrefe, I. Akin, M. Sigl, Coprevalence and incidence of lung cancer in patients screened for abdominal aortic aneurysm, Anti-cancer Research 40 (7) (2020) 4137–4145. doi:10.21873/anticanres.14413.

[29] B. Wiles, M. Comito, N. Labropoulos, L. A. Santore, T. Bilfinger, High prevalence of abdominal aortic aneurysms in patients with lung cancer, Journal of Vascular Surgery 73 (3) (2021) 850–855. doi:10.1016/j.jvs.2020.05.069.

[30] K. I. Alnahhal, V. Urhiafe, M. Narayanan, A. Irshad, P. Salehi, Prevalence of abdominal aortic aneurysms in patients with lung cancer, Journal of Vascular Surgery 75 (5) (2022) 1577–1582.e1. doi:10.1016/j.jvs.2021.09.037.

[31] E. Pasqui, L. Luzzi, E. Lazzeri, G. Casilli, G. Ferrante, C. Catelli, P. Paladini, G. de Donato, Prevalence of concomitant aortic disease and lung cancer: an exploratory study, Journal of Thoracic Disease 16 (5) (2024) 2800–2810. doi:10.21037/jtd-23-1547.

[32] H. R. Gwon, A. Woo, S. H. Yong, Y. M. Park, S. Y. Kim, E. Y. Kim, J. Y. Jung, Y. A. Kang, M. S. Park, D. Y. Kang, S. Y. Park, S. H. Lee, J. S. Kwon, Cross-sectional study of lung cancer patients as a potential high-risk factor for abdominal aortic aneurysm, PLoS ONE 20 (1) (2025) e0315898. doi:10.1371/journal.pone.0315898.

[33] G. M. Stella, F. R. Bertuccio, C. Novy, C. Bortolotto, I. Salzillo, F. Perrotta, V. D’Agnano, V. Conio, V. Arici, P. Cerveri, A. Bianco, A. G. Corsico, A. Bozzani, From copd to smoke-related arteriopathy: The mechanical and immune-inflammatory landscape underlying lung cancer distant spreading–a narrative review, Cells 14 (16) (2025) 1225. doi:10.3390/cells14161225.

[34] D. Costa, M. Andreucci, N. Ielapi, G. F. Serraino, P. Mastroroberto, U. M. Bracale, R. Serra, Vascular biology of arterial aneurysms, Annals of Vascular Surgery 94 (2023) 378–389. doi:10.1016/j.avsg.2023.04.008.

[35] J. J. M. Vermeulen, M. Meijer, F. B. G. de Vries, M. M. P. J. Reijnen, S. Holewijn, D. H. J. Thijssen, A systematic review summarizing local vascular characteristics of aneurysm wall to predict progression and rupture risk of abdominal aortic aneurysms, Journal of Vascular Surgery 77 (1) (2023) 288–298.e2. doi:10.1016/j.jvs.2022.07.008.

[36] A. C. Márquez-Sánchez, E. K. Koltsova, Immune and inflammatory mechanisms of abdominal aortic aneurysm, Frontiers in Immunology 13 (2022) 989933. doi:10.3389/fimmu.2022.989933.

[37] A. Coclite, G. M. Coclite, D. De Tommasi, Capsules rheology in carreau-yasuda fluids, Nanomaterials 10 (11) (2020) 2190. doi:10.3390/nano10112190.

[38] J. Chen, X. Y. Lu, Numerical investigation of the non-newtonian pulsatile blood flow in a bifurcation model with a non-planar branch, Journal of Biomechanics 39 (5) (2006) 818–832. doi:10.1016/j.jbiomech.2005.02.003.

[39] M. M. Khan, R. R. Puniyani, N. G. Huilgol, M. A. Hussain, G. G. Ranade, Hemorheological profiles in cancer patients, Clinical Hemorheology and Microcirculation 15 (1) (1995) 37–44.

[40] D.-e. Hu, J.-c. Ruan, P.-q. Wang, Hemorheological changes in cancer, Clinical Hemorheology and Microcirculation 8 (6) (1988) 945–956.

[41] L. Dintenfass, Haemorheology of cancer metastases: An example of malignant melanoma. survival times and abnormality of blood viscosity factors, Clinical Hemorheology and Microcirculation 2 (4) (1982) 259–271.

[42] G. F. von Tempelhoff, L. Heilmann, G. Hommel, K. Pollow, Impact of rheological variables in cancer, Seminars in Thrombosis and Hemostasis 29 (5) (2003) 499–513. doi:10.1055/s-2003-44641.

[43] J. W. Han, P. S. Sung, J. W. Jang, J. Y. Choi, S. K. Yoon, Whole blood viscosity is associated with extrahepatic metastases and survival in patients with hepatocellular carcinoma, PLoS ONE 16 (12) (2021) e0260311. doi:10.1371/journal.pone.0260311.

[44] V. Tavares, J. Savva-Bordalo, M. Rei, J. Liz-Pimenta, J. Assis, D. Pereira, R. Medeiros, Haemostatic gene expression in cancer-related immunothrombosis: Contribution for ve-nous thromboembolism and ovarian tumour behaviour, Cancers 16 (13) (2024) 2356. doi:10.3390/cancers16132356.

[45] P. Veluswamy, M. Wacker, M. Scherner, J. Wippermann, Delicate role of pd-l1/pd-1 axis in blood vessel inflammatory diseases: Current insight and future significance, International Journal of Molecular Sciences 21 (21) (2020) 8159. doi:10.3390/ijms21218159.

[46] P. Sun, L. Zhang, Y. Gu, S. Wei, Z. Wang, M. Li, W. Wang, Z. Wang, H. Bai, Immune checkpoint programmed death-1 mediates abdominal aortic aneurysm and pseudoaneurysm progression, Biomedicine & Pharmacotherapy 142 (2021) 111955. doi:10.1016/j.biopha.2021.111955.

[47] H. Bai, Z. Wang, M. Li, P. Sun, S. Wei, W. Wang, Z. Wang, Y. Xing, J. Li, A. Dardik, Inhibition of programmed death-1 decreases neointimal hyperplasia after patch angioplasty, Journal of Biomedical Materials Research Part B: Applied Biomaterials 109 (2) (2021) 269–278. doi:10.1002/jbm.b.34698.

[48] A. J. Lee, F. G. Fowkes, M. N. Carson, G. C. Leng, P. L. Allan, Smoking, atherosclerosis and risk of abdominal aortic aneurysm, European Heart Journal 18 (4) (1997) 671–676. doi:10.1093/oxfordjournals.eurheartj.a015314.

[49] G. Danaei, S. Vander Hoorn, A. D. Lopez, C. J. Murray, M. Ezzati, Causes of cancer in the world: comparative risk assessment of nine behavioural and environmental risk factors, The Lancet 366 (2005) 1784–1793.

[50] B. Moorthy, C. Chu, D. J. Carlin, Polycyclic aromatic hydrocarbons: from metabolism to lung cancer, Toxicological Sciences 145 (1) (2015) 5–15. doi:10.1093/toxsci/kfv040.

[51] R. Goldman, L. Enewold, E. Pellizzari, J. B. Beach, E. D. Bowman, S. S. Krishnan, P. G. Shields, Smoking increases carcinogenic polycyclic aromatic hydrocarbons in human lung tissue, Cancer Research 61 (17) (2001) 6367–6371.

